# Associations Between Wearable-Derived Sleep and Physiological Metrics With Performance in Professional Golfers

**DOI:** 10.1101/2025.03.31.25324953

**Authors:** Gregory J. Grosicki, William von Hippel, Finnbar Fielding, Jeongeun Kim, Christopher Chapman, Kristen E. Holmes

**Affiliations:** Performance Science, WHOOP Inc., Boston, Massachusetts, United States; Research with Impact, Brisbane, Queensland, Australia

**Keywords:** wearable technology, sleep consistency, heart rate variability, resting heart rate, athlete monitoring

## Abstract

**Purpose:** Consistently performing at the highest level in golf requires a complex interplay of physiological and psychological attributes, with success often defined by razor-thin margins. Sleep characteristics and cardiac autonomic function, reflected by resting heart rate (RHR) and heart rate variability (HRV), are key indicators of recovery and readiness to perform. Yet, their relevance to elite golf performance remains largely unexplored.

**Methods:** We analyzed wearable-derived longitudinal data from 389 professional tour-level golfers across 521 competitive events (2017-2025), encompassing 35,140 nights of sleep and biometric monitoring. Key metrics included sleep duration (7.2±0.7hrs), sleep consistency (69.1*±*6.9%), RHR (55.9*±*7.9bpm), HRV (64.2±28.1ms), and a composite Recovery score (59.1±9.9%). Golf performance (total score, great shots, poor shots, strokes gained) was extracted from a subscription-based database. Linear mixed-effects models assessed both between-person differences and within-person season-to-season changes, adjusting for age (34.1*±*9.1yrs), height (1.81*±*0.07m), and weight (83.2*±*10.6kg).

**Results:** Golfers with superior sleep and biometric profiles consistently performed better, both between and within individuals (Ps<0.05). Between individuals, each additional hour of sleep was associated with a lower score (b=-0.522), as was a 10-percentage point increase in sleep consistency (b=-0.382), a 1bpm lower RHR (b=-0.038), and a 10-percentage point increase in Recovery (b=-0.476). Within athletes, season-to-season improvements in sleep consistency (b=-0.193 per 10-percentage points), HRV (b=-0.016 per 1ms), and Recovery (b=-0.238 per 10 percentage points) were also associated with lower scores (Ps<0.05).

**Conclusions:** Sleep and measures of cardiac autonomic function are associated with performance in elite golf. Both individual differences and within-athlete improvements were linked to success, highlighting the potential role of sleep, resting heart rate, and heart rate variability in optimizing performance at the highest level of sport.

## INTRODUCTION

In the arena of elite performance, the difference between victory and defeat is often razor thin. Over the last decade, three Masters Tournament champions were decided by just a single stroke, with the 2017 event culminating in a sudden-death playoff – a testament to the fine margins that define success in professional golf. Given this reality, athletes and coaches have increasingly turned to strategies focused on “marginal gains”, where small, incremental improvements translate into meaningful competitive advantages (1). Consistently performing at the highest level in golf requires a complex interplay of physiological and psychological attributes, including motor control, emotional regulation, and cognitive resilience (2, 3). By appreciating these unique performance demands and systematically monitoring training load and readiness, via both subjective inputs and objective indicators such as sleep quality and cardiac autonomic responses, athletes and their support teams can optimize performance and capitalize on these critical margins (4).

Sleep and measures of cardiac autonomic function, reflected by resting heart rate (RHR) and heart rate variability (HRV), have emerged as key indicators of both recovery and readiness to perform. Across numerous sporting disciplines, these metrics have been linked to injury risk (5), training adaptation (6), and cognitive processing (7), highlighting their essential role in sustained high-level performance. Yet, despite the clear performance relevance of these measures, how sleep and autonomic balance influence elite golf performance remains almost entirely unexplored. It may be assumed that elite athletes, particularly golfers who rely heavily on technical precision and mental acuity (8), are already fully optimized in areas like sleep and autonomic regulation. However, if even minor improvements in sleep or autonomic balance can enhance recovery, sustain focus, or support physiological stability, then their role in elite golf performance may be undervalued.

The current research explores this possibility by analyzing over 35,000 nights of longitudinal data collected from nearly 400 professional tour-level golfers across more than 500 elite-level events. We hypothesized that golfers with more favorable sleep and biometric profiles would demonstrate superior performance outcomes.

## MATERIALS & METHODS

### Participants

Participants consented to the use of their anonymized data for research purposes. We analyzed biometric, sleep, and performance data from 389 professional tour-level golfers. Daily data were collected from participants between 2017 and 2025, with participants contributing an average of 3.3 ± 1.8 years of data (range: 1 to 8 years). A total of 521 competitive events were included. For each tournament round, we extracted data from the night immediately preceding play, resulting in 35,140 total nights of data analyzed. These included nights before each competitive round across all four days of tournament play, regardless of whether a participant made the cut. Because all WHOOP data were confidential and securely stored, this study was deemed exempt from Institutional Review Board (IRB) oversight by Salus’s IRB (#6483).

### Data Collection

Daily biometric and sleep data from each athlete were collected using a wrist-worn wearable device (WHOOP Inc., Boston, MA). The WHOOP strap continuously records heart rate via photoplethysmography and movement via three-axis accelerometer. Key cardiovascular metrics, RHR and HRV, were extracted as a weighted average from the primary sleep episode (9, 10). HRV was calculated using the root mean squared of successive differences (RMSSD) (11). Sleep metrics of interest included total sleep duration and sleep consistency, characterized as regularity of sleep onset and offset times over a 4-day window, with greater weighting of more recent days (12). WHOOP cardiovascular and sleep measures have been validated against gold-standard electrocardiogram and polysomnography, demonstrating a low degree of bias and low precision errors (e.g., <20 min bias and precision errors for sleep duration) (13, 14). An exploratory measure included WHOOP Recovery, an integrated metric of nighttime measures of RHR, HRV, respiratory rate, and sleep duration made relative to each athlete’s historical data (15).

Golf performance data for each athlete were extracted from the DataGolf service (https://datagolf.com/), a subscription-based database that provides comprehensive, real-time historical data on professional golf performance, player statistics, and tournament outcomes. Our primary outcome of interest was overall score, defined as the total number of strokes taken during a competitive round, where fewer strokes indicate better performance. Secondary outcomes of interest included: i) Great Shots: defined as shots that gained ≥0.5 strokes relative to the field; ii) Poor Shots: defined as strokes that lost ≥0.5 strokes relative to the field; iii) Strokes Gained: a composite metric that quantifies a player’s performance relative to the field across all aspects of the game, where higher values indicate better performance; iv) Strokes Gained Tee-to-Green: which isolates performance on all shots excluding putting, where higher values indicate better performance; v) Strokes Gained Putting: which captures performance specifically on the green, where higher values indicate better performance.

### Statistical Analysis

Statistical analyses were conducted using R (version 4.4.2). Golfer descriptive characteristics are presented as means ± standard deviation (SD), and normality was assessed using the Shapiro-Wilk test and visual inspection of quantile-quantile (Q-Q) plots. Individual golfer biometric, sleep, and performance data were aggregated by touring season to reduce noise in event-to-event variability (e.g., weather, course difficulty, travel effects) and capture more stable, trait-like physiological patterns. To ensure data quality, all predictors (i.e., sleep and biometrics) and outcome variables (i.e., golf performance) were assessed for potential outliers using the interquartile range (IQR) method (9). Outliers were defined as values exceeding quartile 3 + 1.5 x IQR or falling below quartile 1 - 1.5 x IQR for a given athlete and season. Because no outliers were identified all available data were retained for analysis.

To examine associations between athlete sleep and biometric data with golf performance, we modeled both between-person (each participant’s average data across all available seasons) and within-person effects (season-to-season changes from their personal average, captured via person-mean centering). Three sets of linear mixed-effects models were estimated: one for sleep variables (duration and consistency), one for biometric variables (RHR and HRV), one for Recovery. All models were adjusted for age, height, and weight. To account for the nested structure of the data (i.e., multiple seasons per golfer), models included random intercepts for participant ID. Initial models also included random slopes for season to allow for individual variability in outcome trajectories. However, these models frequently failed to converge and showed evidence of singular fits, indicating that the data did not support the added complexity. As a result, random intercept-only models were used, which sufficiently accounted for within-person variability without overfitting. Both unstandardized (b) and standardized (β) regression coefficients were computed, with the former representing the raw unit change in performance outcomes per unit change in the predictor, and the latter expressing effect size in standard deviation units. Model residuals were visually inspected for normality and homoscedasticity using Q-Q and residuals vs. fitted plots, with no major violations observed.

For visualization and group comparison purposes, between-person effects were categorized into tertiles based on participant-level averages. Group differences across tertiles were tested using linear mixed-effects models with a random intercept for participant ID to account for repeated measures, and fixed effects of age, weight, and height. Post hoc comparisons were conducted using Tukey-adjusted pairwise tests via estimated marginal means. Within-person effects were categorized as “increased” or “decreased” based on whether sleep, biometric, or Recovery metrics deviated positively or negatively from each participant’s mean. These change-based groups were compared using linear mixed models with the same covariates and random intercept structure to account for within-subject variation.

Effect sizes (Cohen’s d) were calculated for post hoc comparisons to quantify the magnitude of group differences. For tertile-based analyses, effect sizes reflect pairwise comparisons between low, medium, and high tertiles. For within-person comparisons, Cohen’s d was calculated for differences between athletes who showed an increase vs. decrease in each metric. Statistical significance was set at α ≤ 0.05 for all analyses. Given the exploratory nature of the study and the correlation among outcome measures, we emphasize effect sizes and consistency of observed patterns across models, rather than strict reliance on P-values.

## RESULTS

### Participant Characteristics

Participants had an average age of 34.1 ± 9.1 years, a height of 1.81 ± 0.07 m, and a weight of 83.2 *±* 10.6 kg. Average resting heart rate was 55.9 ± 7.9 bpm, and HRV was 64.2 ± 28.1 ms. Participants averaged 7.21 ± 0.73 hours of sleep per night with a sleep consistency of 69.1 ± 6.9%. Mean Recovery score was 59.1 ± 9.9%.

### Summary of Continuous Model Estimates for Golf Performance

Unstandardized model estimates for continuous variable analyses examining associations between sleep and biometric data with golf performance metrics are provided in **Table 1**. Between-person and within-person effects were directionally similar, but between-person effects were generally greater in number and stronger in magnitude.

**Table 1.** Unstandardized model estimates for predicting golf performance using sleep and biometric data.

|  |
| --- |
| **P < 0.001** |
| **0.001 ≤ P < 0.01** |
| **0.01 ≤ P < 0.05** |
| **0.05 ≤ P < 0.10** |
| **P ≥ 0.10** |

|  | Score | Great Shots | Poor Shots | Strokes Gained | Strokes Gained:<br>Tee-to-Green | Strokes Gained:<br>Putting |
| --- | --- | --- | --- | --- | --- | --- |
| *Between Person* |  |  |  |  |  |  |
| Sleep Duration (Hour) | -0.522 | -0.002 | -0.465 | 0.523 | 0.419 | 0.280 |
| Sleep Consistency (%) | -0.038 | 0.023 | -0.026 | 0.037 | 0.022 | 0.016 |
| Resting Heart Rate (bpm) | 0.038 | -0.001 | 0.031 | -0.021 | -0.026 | -0.021 |
| Heart Rate Variability (ms) | 0.001 | 0.003 | 0.003 | 0.002 | 0.001 | -0.001 |
| Recovery Score (%) | -0.048 | 0.001 | -0.029 | 0.050 | 0.032 | 0.018 |
| *Within Person* |  |  |  |  |  |  |
| Sleep Duration (Hour) | 0.092 | -0.059 | -0.097 | -0.111 | -0.011 | -0.096 |
| Sleep Consistency (%) | -0.019 | 0.004 | -0.016 | 0.020 | 0.005 | 0.005 |
| Resting Heart Rate (bpm) | -0.005 | 0.005 | -0.010 | -0.026 | -0.036 | 0.018 |
| Heart Rate Variability (ms) | -0.016 | 0.002 | -0.005 | 0.011 | 0.010 | 0.001 |
| Recovery Score (%) | -0.024 | 0.011 | -0.022 | 0.022 | 0.025 | 0.009 |

Unstandardized beta coefficients from three separate linear mixed-effects models: sleep (duration and consistency), biometrics (resting heart rate and heart rate variability), and Recovery. Models included fixed effects for both between person comparisons (based on each golfer's average across seasons) as well as within-person changes (season-to-season deviations via person-mean centering). All models adjusted for age, weight, and height, with random intercepts for participant.

At the between-person level, longer and more consistent sleep, lower RHR, and higher Recovery scores were associated with improved performance, including lower overall score, fewer poor shots, and more strokes gained (Ps<0.05). Specifically, each additional hour of sleep was associated with a 0.522-point decrease in score (b=-0.522, β=-0.185), while a 10-percentage point increase in sleep consistency corresponded to a 0.382-point decrease in score (b=-0.382, β=-0.134). Additionally, a 1 bpm lower RHR was linked to a 0.038-point decrease in score (b=-0.038, β=-0.096), and a 10-percentage point increase in Recovery was associated with a 0.476-point lower score (b=-0.476, β=-0.232).

At the within-person level, fluctuations in sleep consistency, HRV, and Recovery Score were also associated with performance changes (Ps<0.05). A 10-percentage point increase in sleep consistency corresponded to a 0.193-point decrease in score (b=-0.193, β=-0.057), while a 1 ms increase in HRV was linked to a 0.016-point decrease in score (b=-0.016, β=-0.079). Likewise, a 10-percentage point increase in Recovery Score corresponded to a 0.238-point decrease in score (b=-0.238, β=-0.082).

Additionally, season-to-season increases in sleep consistency were associated with fewer poor shots and more strokes gained (Ps<0.05). Increases in HRV were also linked to higher strokes gained, while reductions in RHR correlated with strokes gained from tee-to-green (Ps<0.05). Among intra-individual predictors, improvements in Recovery were the strongest indicator of performance improvements, as higher Recovery scores were associated with lower overall scores, fewer poor shots, and more strokes gained overall and from tee-to-green (Ps<0.05).

### Sleep Duration and Consistency Positively Associate with Performance Across Athletes

Categorical between-person comparisons showed greater overall performance, reflected by lower total score, among golfers in the highest sleep duration (>7.51 hrs/night, Cohen’s d = 0.318) and sleep consistency tertiles (>72.1%, Cohen’s d = 0.256) compared to those in the lowest tertiles, who averaged <7.07 hrs of sleep per night and had a sleep consistency of <66.7% (**Figure 1**; Ps<0.01). Moreover, athletes in the highest sleep duration tertile gained more overall strokes (Ps<0.05) than those in either the medium (Cohen’s d = 0.115) or low tertiles (Cohen’s d = 0.334), which appeared to be driven by fewer poor shots and better putting performance than athletes in the lowest tertile (Ps<0.05, Cohen’s d = 0.176-0.235). Similarly, athletes in the high sleep consistency tertile outperformed those in the lower tertile on overall strokes gained (P<0.05, Cohen’s d = 0.259). However, within-person categorical comparisons, which grouped seasons based on intra-individual increases or decreases in sleep duration or consistency, revealed no significant differences in golf performance (**Figure 2**; Ps≥0.05).

**Figure 1.**
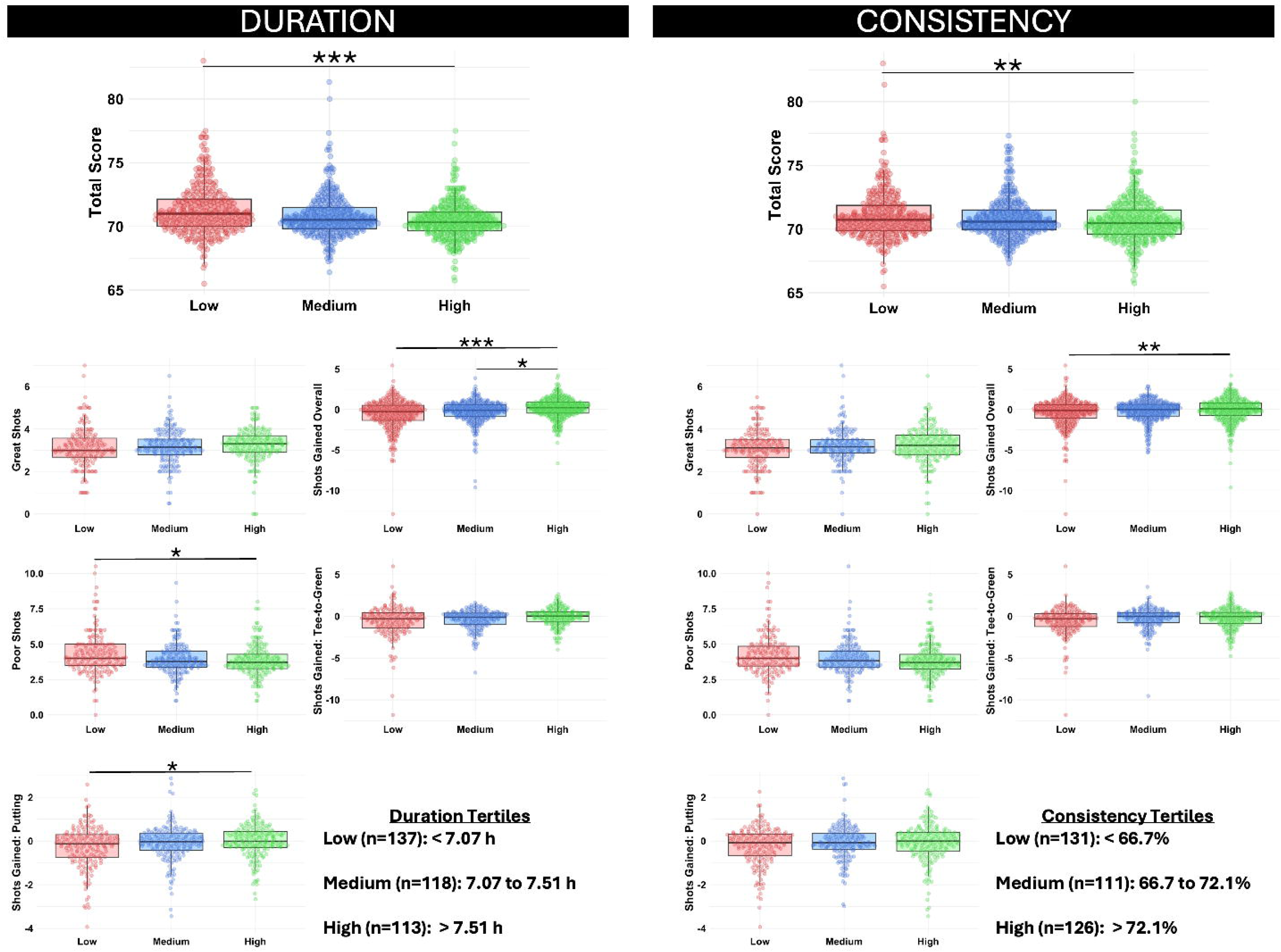
Between-person associations of sleep duration (left) and sleep consistency (right) with golf performance outcomes. Athletes were grouped into tertiles based on their average nightly sleep duration and sleep consistency during their involvement in the study. Swarm plots show distributions for each tertile while boxplots show median and interquartile ranges. Group differences were tested using linear mixed-effect models adjusted for age, weight, and height, with a random intercept for participant ID. When a significant main effect was detected, Tukey-adjusted pairwise comparisons were conducted to identify differences between tertiles. Asterisks indicate statistical significance for these comparisons: *P<0.05, **P<0.01, ***P<0.001.

**Figure 2.**
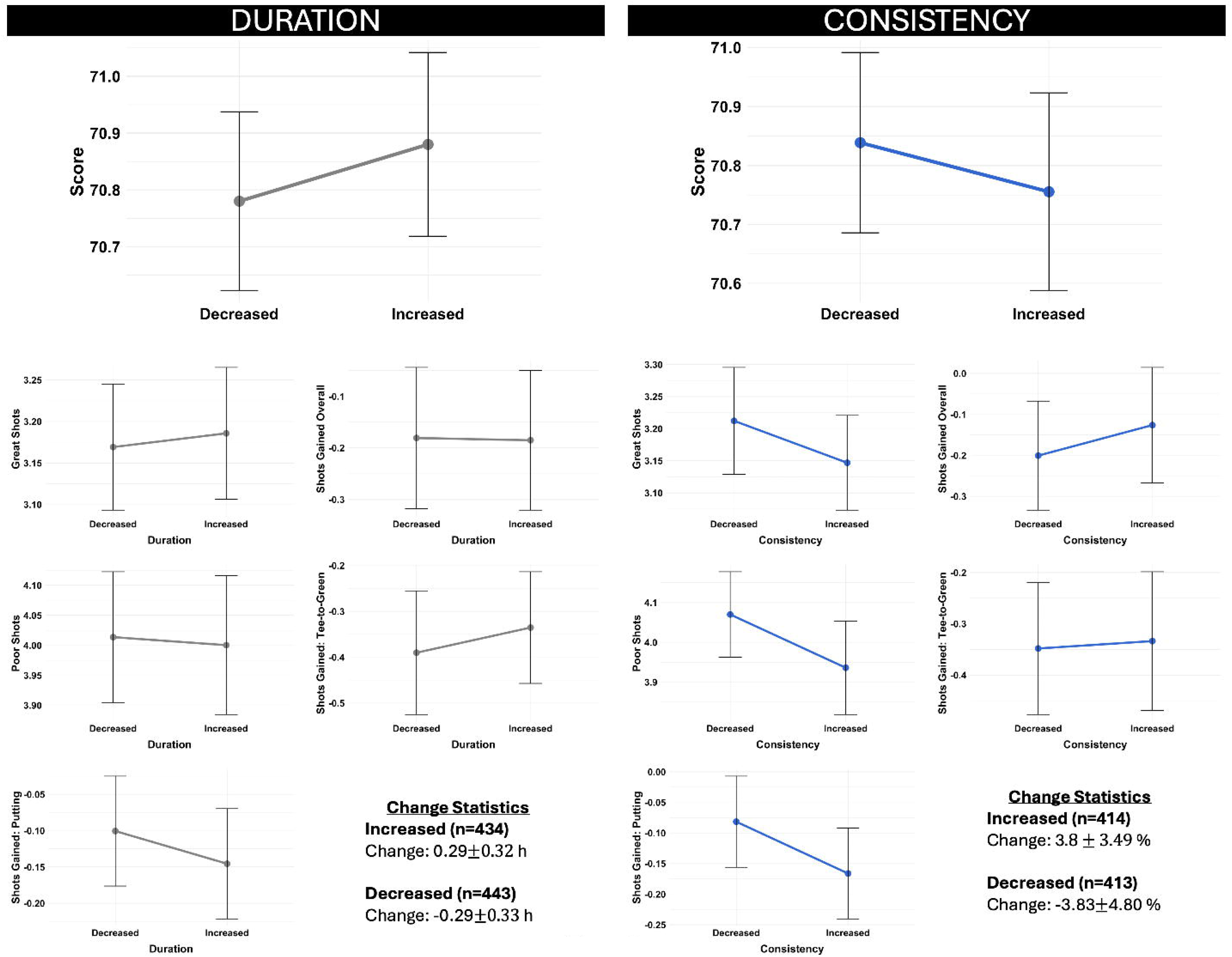
Within-person associations of sleep duration (left) and sleep consistency (right) with golf performance. Years were categorized based on whether sleep metrics increased or decreased relative to each participant’s individual mean. Plotted values represent means and 95% confidence intervals calculated from group-specific standard errors. Group differences were tested using linear mixed-effect models adjusted for age, weight, and height, with a random intercept for participant ID. No asterisks are shown, as no contrasts reached statistical significance (P>0.05).

### Biometric Indicators Show Mixed Associations with Performance

Categorical between-person comparisons showed no significant differences in total score among tertiles (**Figure 3**; Ps≥0.05). However, athletes in the lowest RHR tertile (<52.3 bpm) gained significantly more strokes going from tee-to-green than those in the highest RHR tertile (P<0.05, Cohen’s d = 0.232). Regarding within-person categorical comparisons, intra-individual increases in HRV (Cohen’s d = 0.170 - 0.188) and reductions in RHR (Cohen’s d = 0.170 - 0.217) were associated with more strokes gained overall, and going from tee-to-green (**Figure 4**; Ps<0.05).

**Figure 3.**
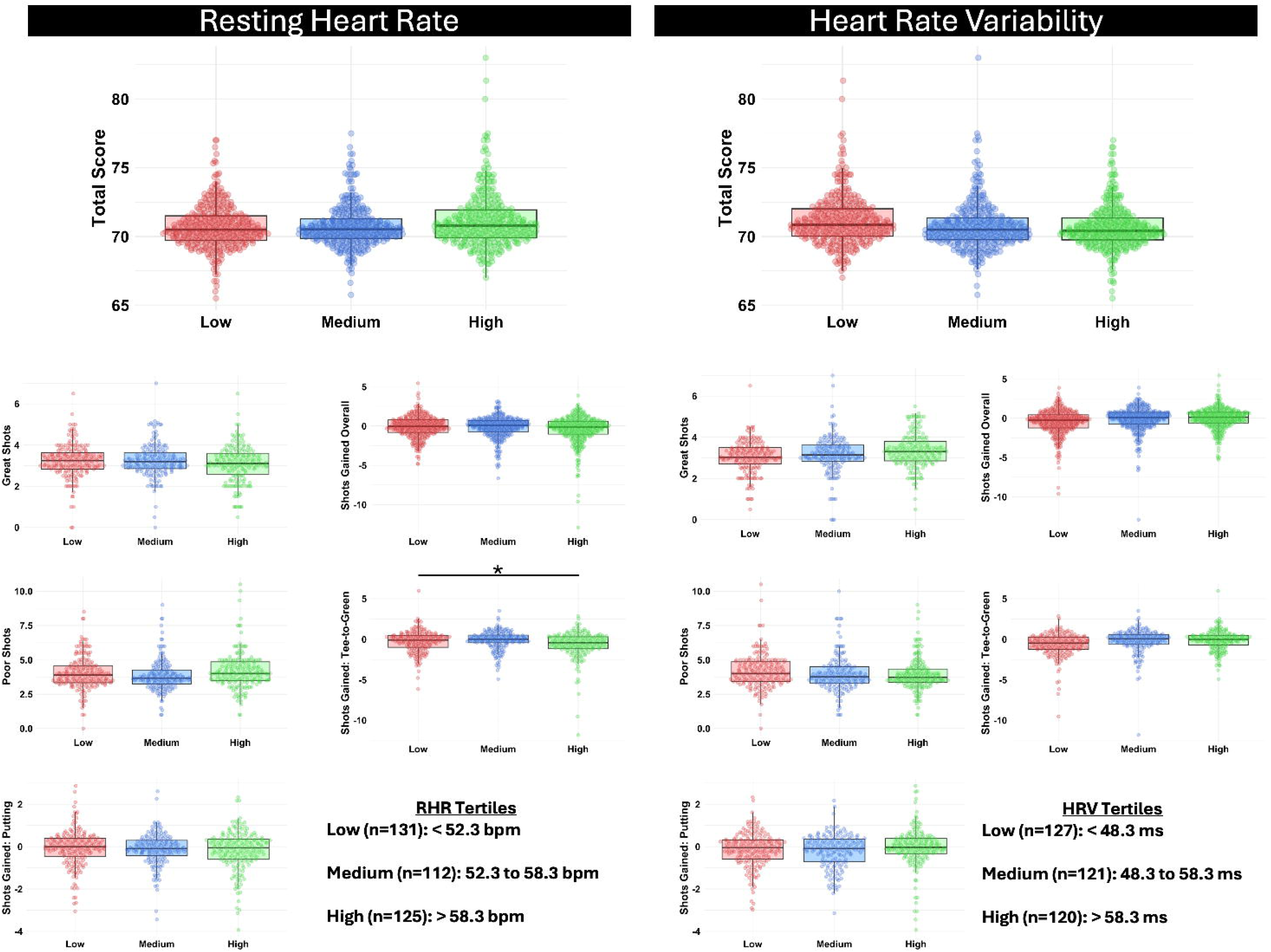
Between-person associations of resting heart rate (RHR; left) and heart rate variability (HRV; right) with golf performance outcomes. Athletes were grouped into tertiles based on their average RHR and HRV during their involvement in the study. Swarm plots show distributions for each tertile while boxplots show median and interquartile ranges. Group differences were tested using linear mixed-effect models adjusted for age, weight, and height, with a random intercept for participant ID. When a significant main effect was detected, Tukey-adjusted pairwise comparisons were conducted to identify differences between tertiles. Asterisks indicate statistical significance for these comparisons: *P<0.05, **P<0.01, ***P<0.001.

**Figure 4.**
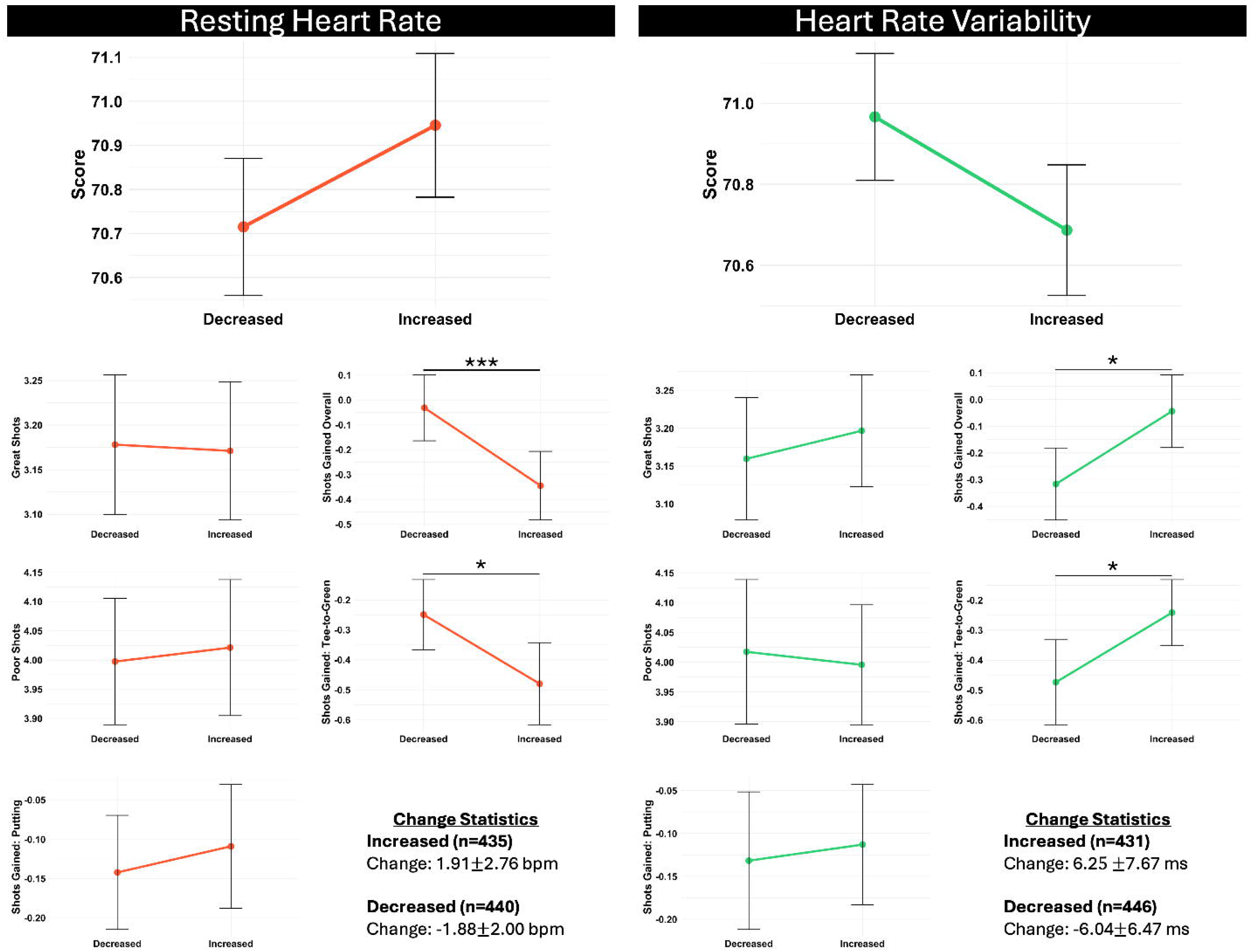
Within-person associations of resting heart rate (RHR; left) and heart rate variability (HRV; right) with golf performance. Years were categorized based on whether biometric variables increased or decreased relative to each participant’s individual mean. Plotted values represent means and 95% confidence intervals calculated from group-specific standard errors. Group differences were tested using linear mixed-effect models adjusted for age, weight, and height, with a random intercept for participant ID. Asterisks indicate statistical significance for these comparisons: *P<0.05, **P<0.01, ***P<0.001.

### Higher Recovery Scores Align with Improved Golf Performance

Categorical between-person comparisons in **Figure 5** showed that golfers in the highest (>65.1%, Cohen’s d = 0.350) and medium Recovery tertiles (59.2 to 65.1%, Cohen’s d = 0.396) demonstrated superior performance, indexed by lower total scores, compared to those in the lowest Recovery tertile (<59.2%; Ps<0.001). Golfers in high and medium Recovery tertiles gained more strokes overall than those in the lowest tertile (Ps<0.001, Cohen’s d = 0.410 – 0.431), likely driven by better (Ps<0.05) performance tee-to-green (Cohen’s d = 0.211 – 0.235) and fewer poor shots (Cohen’s d = 0.242 – 0.269). At the within-person level, years in which an individual’s Recovery increased were associated with better overall performance, reflected by lower total score (**Figure 6**; Ps<0.05, Cohen’s d = 0.120), presumably due to an increased number of great shots (Cohen’s d = 0.213) and more strokes gained (Ps<0.05), both overall (Cohen’s d = 0.159) and on putting (Cohen’s d = 0.164).

**Figure 5.**
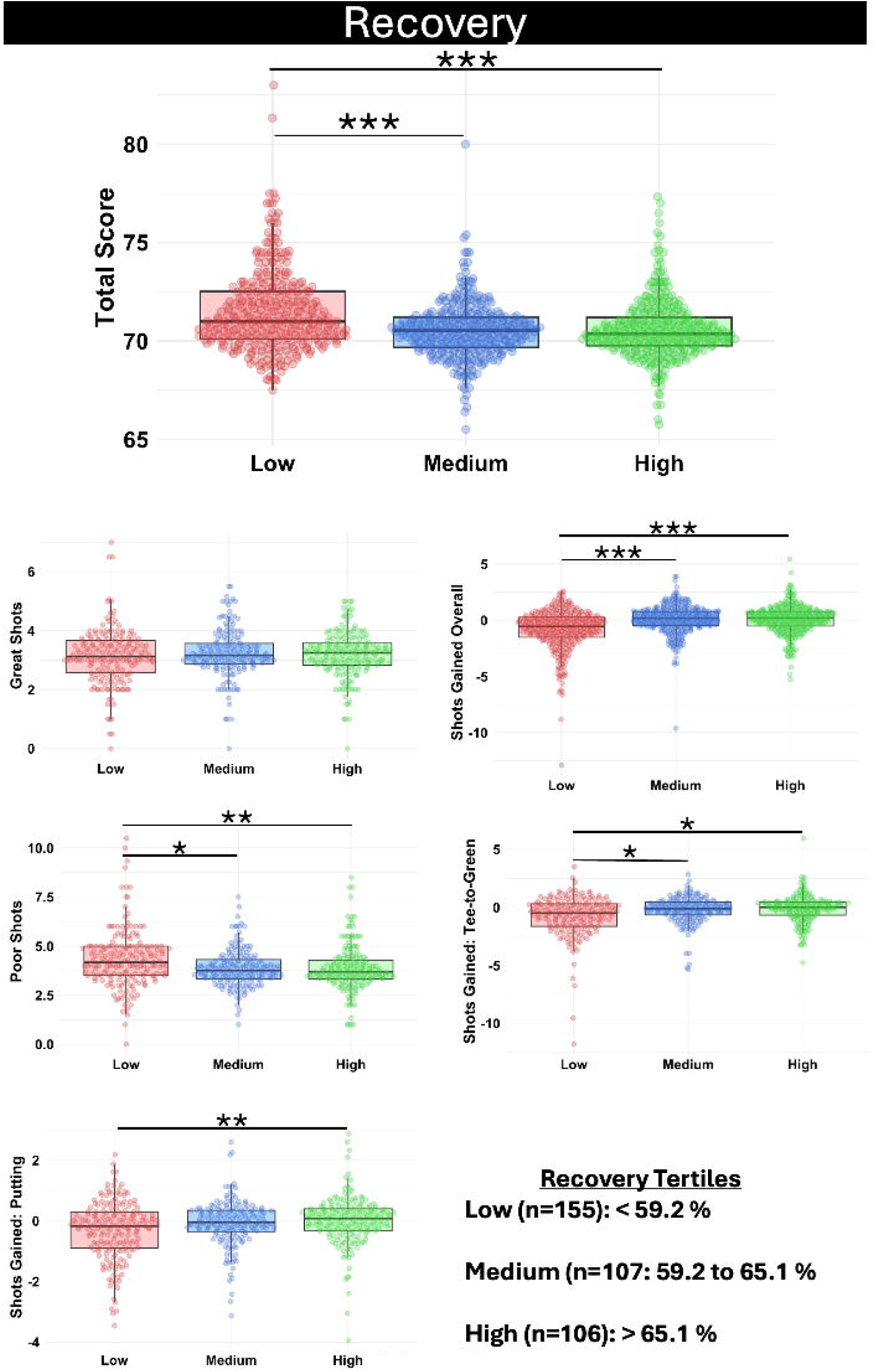
Between-person associations of Recovery with golf performance outcomes. Athletes were grouped into tertiles based on their average Recovery during their involvement in the study. Swarm plots show distributions for each tertile while boxplots show median and interquartile ranges. Group differences were tested using linear mixed-effect models adjusted for age, weight, and height, with a random intercept for participant ID. When a significant main effect was detected, Tukey-adjusted pairwise comparisons were conducted to identify differences between tertiles. Asterisks indicate statistical significance for these comparisons: *P<0.05, **P<0.01, ***P<0.001.

**Figure 6.**
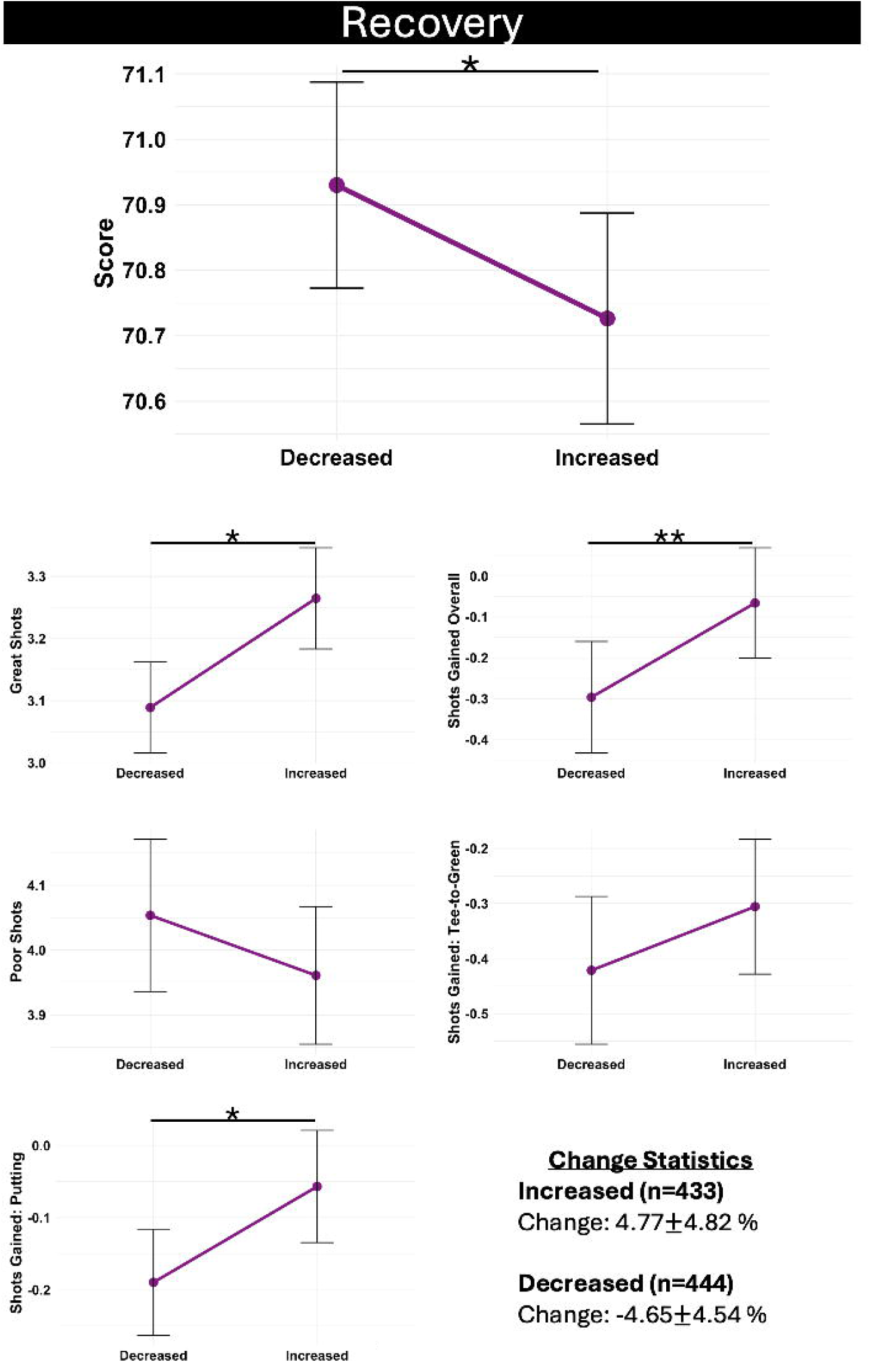
Within-person associations of Recovery with golf performance. Years were categorized based on whether Recovery increased or decreased relative to each participant’s individual mean. Plotted values represent means and 95% confidence intervals calculated from group-specific standard errors. Group differences were tested using linear mixed-effect models adjusted for age, weight, and height, with a random intercept for participant ID. Asterisks indicate statistical significance for these comparisons: *P<0.05, **P<0.01, ***P<0.001.

## DISCUSSION

Leveraging an unprecedented dataset from nearly 400 professional tour-level golfers, we examined whether sleep characteristics and measures of cardiac autonomic function were associated with elite golf performance. The longitudinal nature of the dataset enabled a robust analytical framework to evaluate both between-athlete differences and within-athlete changes across touring seasons. As hypothesized, golfers with more favorable sleep and biometric profiles tended to perform better in competition. Moreover, year-to-year improvements in these metrics within individual athletes forecasted performance gains.

Golfers who habitually demonstrated superior sleep and cardiac autonomic profiles performed at a higher level, evidenced by better overall scores, fewer poor shots, and more strokes gained from tee-to-green and while putting. Sleep is a fundamental biological process that plays a critical role in training, recovery, and performance in elite athletes (16). To our knowledge, we are the first to demonstrate associations between both sleep duration and sleep consistency with objective measures of golf performance. However, two small scale studies have highlighted the potential relevance of sleep in this context. In collegiate golfers, sleep restriction was shown to negatively affect self-reported putting ability (17), while treatment of obstructive sleep apnea with nasal positive airway pressure improved performance in recreational middle-aged golfers (55±9.2 yrs; initial handicap: 12.4 ± 3.1) (18). Although cardiac autonomic responses were not reported in that study, improvements in overnight RHR and HRV would be expected following such therapy (19).

While prior studies have characterized cardiac autonomic responses during golf (20–22), few have examined whether these resting physiological markers relate to competitive outcomes. One small study reported HRV differences between golfers with different handicaps but found no association between HRV reactivity during putting and performance (23). In our data, we observed no between-person associations between HRV and performance. However, RHR was consistently linked with better play, with categorical models highlighting a particularly strong influence on tee-to-green performance. Although speculative, the lower RHR observed among high-performing golfers may reflect greater overall fitness, which has been linked to better golf performance including lower overall scores and enhanced performance from the tee and in the fairway (24, 25). Taken together, these findings suggest that habitual physiological readiness, especially when marked by sleep of sufficient duration and consistency, along with a lower RHR, may confer a performance advantage at the highest level of golf.

Consistent with group-level trends, within-individual improvements in sleep and biometric data were associated with better golf performance, further alluding to the potential value of optimizing these metrics. Notably, while no benefits were observed with sleep extension (i.e., increasing sleep duration), enhancements in sleep consistency were linked with lower overall scores, fewer poor shots, and more strokes gained. Sleep extension is an emerging and ecologically relevant intervention with potential to enhance performance and healthspan (26). However, evidence supporting performance improvements with sleep extension is limited, and outcomes appear to be highly context-dependent (27). Our real-world study adds to this small but growing body of literature by showing that, in a large cohort of elite golfers, increasing sleep duration did not translate to performance improvements. However, because baseline sleep duration already exceeded the ∼7 hours per night associated with the lowest risk for mortality (28), the potential for additional gains may have been limited. Meanwhile, the benefits of sleep consistency in our study align with prior findings in collegiate soft tennis players, where greater variability in sleep duration was inversely associated with service score (29). Additionally, among elite team sport athletes, greater sleep consistency has been linked to improved sleep efficiency (30), further underscoring the value of minimizing variation in sleep-wake timing. Together with emerging evidence suggesting that sleep consistency may be a stronger predictor of mortality than sleep duration alone (31), our findings highlight the need for future experimental studies exploring whether optimizing sleep consistency can enhance athletic performance.

Within a given athlete, improvements in HRV were associated with lower overall scores, while reductions in RHR and increases in HRV were linked to strokes gained, likely driven by superior tee-to-green performance. These findings align with prior research linking rising HRV trends to favorable adaptations and improvements in overall fitness (6, 32). However, unlike much of the previous research in this area that has relied on post-waking HRV measures (33–35), we measured HRV during sleep. While morning and nocturnal measures of RHR and HRV are moderate-to-highly correlated (6, 36, 37), recent research in recreational runners suggests that nocturnal HRV is more responsive to physical loading and associated with positive training adaptations (6). However, a potential limitation of relying solely on nocturnal HRV as a readiness marker is HRV saturation (38), a phenomenon whereby HRV decreases independently of fatigue and coincides with decreases in RHR. This is postulated to result from acetylcholine receptor saturation at the cardiac myocyte level, leading to sustained parasympathetic control of the sinus node and diminished respiratory heart modulation (32, 39). Notably, a RHR of 60 bpm has been identified as a HRV saturation “tipping point” (32), below which interpretation may be confounded. In the context of the present study, where average RHR was ∼56 bpm, this may partially explain the lack of significant between-person associations between HRV and performance, despite consistent associations with RHR. Future work may benefit from supplementing nocturnal data collection with post-waking assessments, particularly those taken in a standing position to introduce an orthostatic challenge and reduce the likelihood of vagal saturation.

An exploratory aim of our study was to examine associations between Recovery, a composite metric encompassing RHR, HRV, respiratory rate, and sleep duration, with golf performance. Notably, Recovery emerged as the single most consistent contributor to performance in within-person models and was comparable to independent sleep and biometric variables when analyzed as a between-person predictor. This finding suggests that aggregating complementary physiological signals into a holistic readiness metric may provide a more informative indicator of performance potential. A key distinction of the Recovery score is that it adapts to individual trends over time, potentially offering a more personalized perspective on readiness compared to static physiological measures. While these features may contribute to the predictive strength of Recovery in our study, caution is warranted in interpreting wearable-derived recovery insights. These algorithms are typically developed using population-level trends and may not fully capture intra-individual variability in training response (40). Moreover, such metrics often exclude subjective dimensions of readiness, such as mental fatigue, effort, stress, and motivation, which are important moderators of the relation between training load, injury risk, and performance (41).

Several strengths and limitations of this study should be acknowledged. First, despite our use of within-person models, the study’s observational design limits causal inference. Unmeasured factors such as training load, travel schedules, nutrition, or psychological stress could influence sleep and physiological metrics and performance, potentially confounding analysis. Second, all data were collected from a single wearable platform (WHOOP), which, while validated (13, 14), may have its own systematic biases or errors and may limit generalizability of study findings. For example, because WHOOP extracts biometrics as a weighted average from the primary sleep episode (9), these values may differ from unweighted nightly averages. Additionally, updates to the device hardware and algorithms over the study period may have influenced recorded values; however, parity was always evaluated before such changes were implemented to ensure data consistency over time. Generalizability may also be limited by the unique nature of our sample. This elite-level, self-selected group is likely more health-conscious than the average golfer, potentially limiting extrapolation to broader athletic or recreational populations. Nonetheless, our use of a large, real-world dataset from nearly 400 professional golfers is also a key strength. Additional strengths include the use of a validated wearable device and the integration of granular, objective performance metrics (e.g., strokes gained, tee-to-green, putting performance, and shot quality counts) directly from competition. Last, to our knowledge, our study is the largest to date examining associations between sleep, measures of cardiac autonomic function, and objective golf performance.

In conclusion, we find that sleep and measures of cardiac autonomic function are associated with performance in elite golf. Importantly, these associations were evident both between and within athletes, where seasonal improvements in these metrics forecasted better play. Together, these findings highlight the value of consistent, high-quality sleep and favorable measures of cardiac autonomic balance as contributors to high-level golf performance.

## ACKNOWLEDGEMENTS

None

## CONFLICTS OF INTEREST

GJG, WvH, FF, JK, CC, and KEH are employees of WHOOP, Inc.

The results of the study are presented clearly, honestly, and without fabrication, falsification, or inappropriate data manipulation. The results of the study do not constitute endorsement by the American College of Sports Medicine.

## DATA AVAILABILITY

The data that support the findings of this study are not publicly available due to intellectual property concerns of WHOOP, Inc. Data may be made available upon request to the company via for researchers who meet the criteria for access to confidential data.

## Abbreviations

HRV: Heart rate variability
RHR: Resting heart rate

## Notes

### Author Declarations

Participants consented to the use of their anonymized data for research purposes. Because all WHOOP data were confidential and securely stored, this study was deemed exempt from Institutional Review Board (IRB) oversight by Salus IRB (6483).

